# Lived experiences and occupational pressures among Vietnamese nail salon workers in Maryland

**DOI:** 10.64898/2026.09.10.26362742

**Authors:** Kevin K. Nguyen, Emily M. Agree

**Author notes:** Correspondence should be addressed to K.K.N. and E.M.A.

## Abstract

Individuals of Vietnamese descent comprise a majority of the United States nail salon workforce. Whereas prior research has centered on chemical exposures, thematic analysis of interviews with Vietnamese nail salon workers around Baltimore, Maryland examined the lived experiences of nail salon workers. Six themes were identified: (1) nail work as a familial livelihood and a sacrifice-oriented pathway towards intergenerational mobility; (2) perceived flexibility within the workplace with low control; (3) professional pride grounded in artistry, detail, and customer care; (4) cumulative occupational risks; (5) emotional labor as an expected and core component of nail salon work; and (6) limited perceived capacity to change working conditions. These findings suggest that the occupational pressures in nail salon work cannot be solely framed as a lack of worker knowledge or motivation towards adopting safer practices. Intervention success depends on a holistic view of workers’ lives, whose choices are shaped by family obligations, customer demands, and conditions largely outside their control. Therefore, efforts to improve nail salon health may benefit from shifting the burden of risk management away from individual workers and engaging the wider community.

## Introduction

The nail salon industry in the United States has expanded rapidly over the past decade. In 2023, the industry generated over $2.8 billion in revenue and is projected to reach $4.3 billion by 2030 [1]. As of 2024, around 210,000 manicurists and pedicurists were employed nationwide [2]. Employment in the sector is projected to grow 7% over the next decade, faster than the average for all occupations (3%) [2]. Most salons are small, family-run businesses with fewer than ten employees, and the workforce is overwhelmingly composed of women (81%) and first-generation immigrants (79%) [3]. Vietnamese-Americans constitute over half of all licensed nail technicians, own a substantial share of salons nation-wide, and have established the nail salon as a defining ethnic economic niche over the past several decades [3–5]. The nail salon industry’s low barriers to entry, including minimal formal education requirements, the ability to work without English fluency, and established ethnic community networks, have made it a primary livelihood for Vietnamese immigrants in particular [4–7].

Despite the ubiquity of nail salons in American communities, the workers who staff them remain among the most overlooked and vulnerable members of the service workforce. Nail technicians are chronically exposed to a complex mixture of chemical hazards present in the products they handle daily. Of particular concern are volatile organic compounds (VOCs) such as toluene, ethyl acetate, methyl methacrylate, and formaldehyde, as well as plasticizers including dibutyl phthalate [8–10]. These exposures have been linked to respiratory and dermal irritations, headaches, cognitive impairment, and adverse reproductive health outcomes [11–17]. Beyond chemical hazards, nail salon workers face ergonomic risks from prolonged work in constrained postures, potential exposure to bloodborne and airborne pathogens, and psychosocial stressors including long work hours, commission-based pay structures, and customer-driven demands [7, 8, 18–21]. Research suggests that the lifetime cancer risk for nail technicians may be substantially elevated [22].

Evidence-based interventions to improve nail salon worker health remain scarce. The few documented intervention studies have been geographically concentrated in certain regions, where academic–community partnerships have developed culturally-tailored training programs for salon owners and workers [23, 24]. These interventions have generally shown promise in improving occupational health knowledge, self-efficacy, and some self-reported safety behaviors. However, implementation research has also revealed persistent barriers, including the high cost and limited availability of safer products, lack of engagement from some salon owners, resistance to certain safety practices, fragmented regulatory enforcement, and the broader economic pressures facing immigrant-owned small businesses [24, 25]. Qualitative studies have been pivotal in uncovering the complex interplay between personal, organizational, and structural factors that shape health and safety practices in this setting [6, 21, 26], yet the voices and lived experiences of workers themselves remain underrepresented in the literature.

Maryland has received limited attention in research on the occupational health experiences of its nail salon workforce. The state’s regulatory environment, local labor markets, demographic composition, and cultural dynamics may differ from regions where prior research has been conducted, such as California, Pennsylvania, New York, and other states. Whereas those regions benefit from established networks of worker-serving organizations and voluntary healthy nail salon recognition programs, Maryland has no comparable program and a smaller, geographically dispersed Vietnamese community that may limit the reach of community-based support. The factors that draw workers into the profession, the conditions that keep them there, and the barriers they face in protecting their health have not been systematically explored in this context. Furthermore, existing studies have tended to focus on occupational exposures rather than examining the broader lived experience of nail salon work, including the profession’s role within the family, its emotional demands, and workers’ perceptions of their own capacity for change.

Knowledge does not always translate into changed behavior. Despite prior nail salon interventions improving what nail salon workers know, changes in salon operations remain inconsistent [23]. This suggests that future programs may depend less on transmitting information and more on understanding the familial obligations, customer relationships, and structural constraints that govern daily salon operations. This study addresses these gaps through a qualitative investigation of the lived experiences of Vietnamese nail salon workers in the Baltimore, Maryland metropolitan area. We conducted semi-structured interviews with Vietnamese nail salon workers and supplemented these with an open-ended survey to identify the factors that compromise health and safety within the salon, characterize barriers to adopting safer practices, and document worker-identified priorities for improvement. Our findings contribute to the growing body of work on occupational health disparities among nail salon workers by centering workers’ own narratives and extending the geographic scope of inquiry to a state where no prior health promotion programs for this workforce exist.

## Methods

### Study design

This study comprised two complementary data collection components: in-person semi-structured interviews and a follow-up online survey. This mixed-method approach allowed us to capture in-depth narrative accounts of workers’ experiences while also gathering data on demographic characteristics, salon environments, safety practices, and health perceptions across the sample.

### Participant recruitment

Recognizing the importance of trust and cultural understanding in connecting with nail salon workers, a population that prior research has identified as hard-to-reach and often wary of outsiders [6], we employed a community-based approach in partnership with the Maryland Vietnamese Mutual Association (MVMA), the oldest nonprofit organization serving the Vietnamese-American community in Maryland. We recruited participants using a snowball sampling approach, consistent with established methods for reaching vulnerable and hard-to-reach populations [27]. Initial contacts were identified through MVMA’s community networks, and additional participants were recruited through referrals from participants during in-person visits to nail salons.

Eligibility criteria included being at least 18 years of age, currently working part-time or full-time as a technician or owner in a nail salon in the Maryland area, and self-identifying as Vietnamese or Vietnamese-American. We recruited both technicians and owners and targeted Vietnamese-owned salons, given the high proportion of Vietnamese immigrants in the nail salon workforce nationally and in Maryland specifically. A total of ten workers were recruited for qualitative interviews across multiple salons, and nine workers completed the supplementary survey. Recruitment continued until thematic saturation was attained in the interview data, as evidenced by the recurrence of consistent themes across transcripts.

### Ethical considerations

This study protocol was reviewed and approved by the Johns Hopkins University Homewood Institutional Review Board (HIRB00019771). Given the sensitive nature of the health-related interview questions and the vulnerability of the study population, several safeguards were implemented. Informed consent was obtained from all participants prior to data collection, participation was voluntary, and participants could withdraw at any time. All participants were compensated with a $20 gift card for their time. Prior to analysis, identifying information was removed from interview transcripts during the transcription process, and participants identified only by numeric codes throughout.

### Interview data collection

Semi-structured interviews were conducted with ten nail salon workers. Interviews were conducted one-on-one at participants’ salons and in the participant’s preferred language (Vietnamese or English). The bilingual lead researcher conducted all interviews and, for sessions conducted in Vietnamese, translated and transcribed the recordings into English for analysis. Each interview lasted approximately 30 to 45 minutes and was audio-recorded with the participant’s consent.

The interview guide was semi-structured and exploratory in nature, with open-ended questions designed to elicit participants’ reflections on their occupational experiences, health concerns, and ideas for change. The guide covered the following broad domains: (a) how participants entered the nail profession and what keeps them in it; (b) descriptions of a typical workday, including hours, pace, and break patterns; (c) perceptions of work-related health risks and symptoms; (d) current protective practices and barriers to safer work; (e) sources of stress and strategies for coping; (f) relationships with customers, coworkers, and salon owners; and (g) views on what changes, if any, would improve the industry. Sample questions included: “How has your experience working as a nail technician been?” “How do you feel your work has influenced your health and well-being?” and “What do you think would be most important to change in the salon to make you feel more healthy?” The guide was iteratively refined after initial interviews.

### Survey data collection

Following each interview, the same participants were invited to complete a brief online survey. Survey responses were not linked to individual interview transcripts. This design gave participants anonymity to report on potential sensitive topics. Furthermore, the survey was administered to provide demographic and workplace information that could contextualize the qualitative findings. The survey was developed in both Vietnamese and English and was administered in the participant’s preferred language. It consisted of items organized into several domains: demographics (gender, age, birthplace, years of U.S. residency, and education); salon environment (number of workstations, ventilation systems, and doors/windows); work characteristics (hours per week, customer volume, and relationship to staff); safety practices (use of personal protective equipment, product avoidance, familiarity with hazardous compounds, availability of safety materials, and break patterns); health status (self-rated health, health patterns, and frequency of health-related thinking); workplace climate (comfort discussing salon issues with management, trust in safety precautions, discussion of health with coworkers, and history of leaving a salon for health reasons); and awareness of external resources (awareness of industry health movements, and state government services). One open-ended item asked: “What else could be done to support you, your family, and your neighbors in achieving better health and well-being?” Survey responses were used descriptively and to corroborate themes from the qualitative interviews.

### Data analysis

Interview transcripts were used for systematic coding and analysis. We employed a thematic analysis approach that combined deductive and inductive strategies [28]. Initial coding began with multiple rounds of open coding on the first several transcripts. The research team met to discuss and reconcile codes and an initial coding scheme was developed. Through an iterative process, focused codes were refined and organized into broader thematic categories. Once a stable coding scheme was established, all transcripts were coded using the finalized scheme, with additional codes added as they emerged and reviewed. Initial codes generated across the ten transcripts were organized into 21 thematic categories. After further refinement, six overarching candidate themes were identified that captured the central patterns across narratives. Illustrative quotations were extracted from the transcripts to demonstrate each theme. Participants are identified by numeric codes to protect confidentiality.

Survey data were analyzed descriptively. Frequencies and proportions were calculated for closed-ended items, and open-ended survey responses were reviewed for consistency with the interview themes. The integration of interview and survey data followed a convergent design in which the two data sources were analyzed independently and then merged during the interpretation phase.

## Results

### Participant characteristics

Ten Vietnamese nail salon workers participated in semi-structured interviews across multiple salons in the greater Baltimore metropolitan area. Participants ranged in age from their mid 20s to their early 60s. The sample included six women and four men. All but one participant were born in Vietnam, and the majority had resided in the United States for more than 15 years. Participants held a range of roles, including working technicians, owners, a salon manager, and a semi-retired worker, with experience in the nail industry ranging from 3 to 30 years. Most participants reported working more than 40 hours per week, with several describing workdays of 10 to 12 hours, six or seven days per week. Educational backgrounds varied, from some primary education to completed postsecondary degrees, though most had attained a high school education. Vietnamese was the primary language spoken at home and at work for the majority of participants, though several reported using both Vietnamese and English.

### Summary of themes

Across the interview and survey data, six overarching themes were identified through thematic analysis: (1) nail work as a familial livelihood and sacrifice-oriented pathway toward intergenerational mobility; (2) perceived flexibility within the workplace with low job control; (3) professional pride rooted in artistry, quality, and customer care; (4) cumulative occupational risks including chemical exposures, ergonomic strain, fatigue, and missed breaks; (5) emotional labor as a core component of nail salon work; and (6) limited perceived capacity towards change. Each theme is presented below with illustrative quotations drawn from the interviews.

### Theme 1: Nail work as a familial livelihood and intergenerational sacrifice

Nearly every participant described entering the nail industry through family or community networks. Relatives who were already established in the profession provided the introduction, training, and sometimes the employment itself. One participant explained that her sister-in-law was already doing nails and, drawn by her own interest in art, she decided to learn. Another described a relative offering him a position at the family salon when he was in between jobs, telling him: “Hey, do you want to work … at my salon? I need help anyway. I’d rather offer it to you than a random person and you kind of need it too” (Participant 1127). For many, nail work was accessible. As one technician stated: “It doesn’t require a lot of formal credentials. Anyone can do it. Especially for new immigrants, they often choose nails because they don’t have to go back to school” (Participant 1241). Together, these observations suggest that salons function as more than workplaces, serving as hubs where family and community members gathered, trained one another, and remained connected.

A striking feature of these narratives was the framing of nail work as a sacrifice made for the next generation. Participants uniformly expressed that they did not want their children to follow them into the profession, instead emphasizing the importance of education and other professional careers. One mother described: “I work for you guys [her children], you guys go to school for me” (Participant 1016). Another participant, who had run a salon seven days a week for two decades with almost no vacations, explained: “I don’t wish for my children to go into this business… In order to succeed you need to work nonstop” (Participant 1251). Participants often took pride in their children’s professions as nurses, police officers, lawyers, and engineers. Parents endure demanding work at the salon so that their children can pursue opportunities that were potentially inaccessible to them. Even an older participant who had returned to part-time salon work after retiring emphasized that her family’s children had all been encouraged to study and pursue other careers.

### Theme 2: Flexibility with low control

Participants frequently cited flexibility as a primary reason for remaining in the nail industry. The ability to set one’s own schedule, take time off to attend to children or family obligations, and adjust working hours to personal needs was described as a meaningful advantage over work with more rigid schedules. One participant recalled: “I used to work in a factory. It was hard to take time off. If my child got sick, I couldn’t just leave. With nails, I can take a half day, run to the doctor, then come back” (Participant 1016). Another, who entered nails while attending school, noted: “Salon work let me clock in and out around my classes unlike factories or American-owned shops with fixed shifts” (Participant 1125). For an older participant who worked only on an as-needed basis, the part-time nature of the work was itself the appeal: “I only go in when they need me… Mostly part-time, usually Fridays and Saturdays” (Participant 1529).

However, this flexibility was tied to the uncertainty of commission-based pay and customer-driven demand, leaving workers with little control over their working conditions. Because income was tied directly to the number of clients served, periods of low traffic generated financial worry. One participant noted: “If you’re in a salon all day and there are no customers, even that can bring stress” (Participant 1016). The competitive landscape brought further demands onto the workers. With many salons in close proximity, technicians felt compelled to accept late-arriving customers rather than risk losing them to a competitor. The same participant described: “We’re closed at 7:30pm and sometimes stay until 9pm… If you don’t [take late customers] they’ll just go down the street and won’t ever come back” (Participant 1016). A salon owner similarly noted that customers effectively controlled technicians’ schedules: “If we don’t take the customer, the nail salon next door will. Thus, the customers control the nail techs” (Participant 1157). The tension between flexibility and uncertainty was summarized by one of the participants: “If you want to maximize income, you start early and finish late. If you value balance… start a little later and leave earlier. Earn to your target and call it a day” (Participant 1058).

### Theme 3: Professional pride in artistry and customer care

Despite the challenges of the work, participants expressed pride in their craft and in the relationships they built with clients. Several described nail work as an art form requiring skill and attention to detail. One participant, who had entered the profession because she “liked drawing and being artistic,” had remained in nails because of the satisfaction of “making things beautiful” (Participant 1113). Quality was treated as non-negotiable. A participant stated: “We can’t compromise on quality. We could also add more services, such as body massage, facial. Make customers feel like they are getting their money’s worth and keep them here” (Participant 1251).

The emotional reward of client satisfaction was a recurrent source of meaning. A participant captured this sentiment vividly: “When I finish and a client feels happy, when their sore or rough feet look nice, that makes me happy too. I still feel useful, like I can contribute something” (Participant 1529). For this participant, continuing to work was linked to maintaining a sense of purpose and dignity: “Older people can feel ashamed if they’re not doing anything. Working helps me feel confident and lively” (Participant 1529). Several participants also expressed broader pride in the Vietnamese community’s contribution to the industry. One noted: “When I came in the ‘90s, Vietnamese techs were number one in this industry… Vietnamese people are no longer dominant, but it’s still a reliable path and we are still masters of this profession” (Participant 1125).

### Theme 4: Cumulative occupational risks

Participants described a range of physical and environmental health risks that accumulated over the course of long careers. Chemical exposures were acknowledged primarily through the proxy of odor: participants recognized that strong-smelling products were potentially harmful. One participant noted a preference for low-odor products, explaining: “There are monomers that have a strong smell and others that don’t. So I prefer to use the low-odor monomers for acrylics because it isn’t as strong” (Participant 1125). Strikingly, when workers were surveyed about their familiarity with common nail product chemical compounds, acetone was the only compound identified by any participant. This suggests a gap in workers’ awareness of the hazardous chemicals present in the products they handle daily.

Ergonomic strain was described as an unavoidable feature of the work. Participants reported chronic shoulder, back, and hand pain from maintaining constrained postures while working on clients’ hands and feet. One participant described the cumulative nature of these risks: “Health-wise it’s cumulative: dust, fumes, posture, shoulders, wrists, back, so masks, ventilation, and ergonomics matter” (Participant 1157). Fatigue from long hours was pervasive. Multiple participants described working 10 to 12 hours per day, six or seven days per week, often without adequate meal breaks. One participant recalled 20 years of operating a salon with “sometimes no breakfast no lunch, eating in the car” (Participant 1251). Another acknowledged that on busy days, “if you don’t take breaks to eat, you’ll be drained” (Participant 1058). The lack of structured break time was confirmed by participants, several of whom described taking breaks only “between customers” or “when it slows,” while one participant reported not taking breaks at all.

Masks and ventilation were mentioned as protective measures, though their use appeared variable. From the survey data, participants uniformly reported always using personal protective equipment. Notably, half of the participants reported that their salons had no personal ventilation devices at their workstations.

Three participants reported having been diagnosed by a physician with health conditions that may be associated with salon work, including respiratory problems, skin problems, and musculoskeletal problems. Three of nine participants also reported having left a salon at some point due to concerns about their health or safety. However, when asked to rate their current health status, most participants described it as “normal” or “good,” and the majority did not perceive recurring negative health patterns. This apparent disconnect between reported exposures and perceived health status may reflect a normalization of occupational risk, a pattern consistent with previous research in nail salon settings [6].

### Theme 5: Emotional labor as core to the profession

Across the interviews, emotional self-regulation emerged as a central and expected component of nail salon work. Participants described the constant need to manage their own emotional responses to difficult customers, interpersonal conflicts with coworkers, and the stress of unpredictable income. One participant stated explicitly: “The job itself isn’t stressful; sometimes our mindset makes it stressful” (Participant 1125). Multiple participants described strategies for managing frustration: stepping outside briefly, breathing exercises, compartmentalizing stress, and maintaining a positive outlook. As one participant put it: “I just try my best and stay positive. I tell myself that if I work hard today, tomorrow will be better, and clients will come back” (Participant 1113).

The emotional demands extended beyond individual coping to encompass the management of interpersonal dynamics within the salon. Participants described navigating competition among coworkers for customers and favoritism from owners in allocating walk-in clients. One participant emphasized the importance of deescalation skills: “Some customers won’t be satisfied… I know how to de-escalate and find a way to rectify things most of the time” (Participant 1127). Several participants had deliberately chosen small salons or two-person operations specifically to minimize interpersonal conflict. One explained: “That’s why we only have the two of us in the salon for 7 years now. We don’t want another person coming in and building that stress” (Participant 1125). The framing of stress as an interpersonal or mindset issue, rather than a structural feature of the work, was pervasive and may influence how workers envision change in the salon.

### Theme 6: Limited perceived capacity towards change

When asked about what changes could improve the industry, participants gave a range of responses. Some were unsure about industry-level change, others described individual or incremental product adjudgments, and one participant indicated specific structural changes. One participant stated: “Change? I don’t really know. It’s difficult. I’ve already been doing nail salons for several years so I just want to retire” (Participant 1113). A typical response was: “Every few years products and techniques change, but the work is the same. New products come, we learn them, that’s it. I don’t see a big industry change needed” (Participant 1016). A common thread through these responses is that shaping the conditions of nail salon work is beyond the workers’ reach.

One salon technician-manager identified structural barriers: “Too many unlicensed operators undercut standards. Regulators don’t have the resources to check evenly; some compliant shops get inspected repeatedly while others never do” (Participant 1157). He also noted that customers exercised disproportionate power in the salon-client relationship, drawing a comparison to the medical profession: “Take for example, you book with a doctor. Would you even consider being late?… Exactly!” (Participant 1157). Yet even this participant acknowledged the difficulty of achieving collective change, observing that within Vietnamese culture, “there is no way that I can change you; you need to change for yourself” (Participant 1157). The structural reforms he identified, specifically licensing enforcement, customer accountability, and professional culture, are ones that nail salon workers or owners could not implement on their own.

The follow up survey echoed these observations. When asked whether they were aware of movements to improve worker health in the nail salon industry, four of nine participants indicated awareness, while five did not or were unsure. Only one participant reported awareness of state government services available for salon workers. When asked what could be done to support their health and well-being, participants noted education about healthcare programs, safety, and money. These are needs that are experienced individually and points to structural gaps in the services available to nail salon workers. Collectively, these observations suggest that nail salon workers have a limited scope for industry-level change and highlight that the problems they identify are ones in which they have no mechanism to address.

## Discussion

Our study examined the lived experiences and occupational pressures of Vietnamese nail salon workers in the Baltimore, Maryland metropolitan area, contributing to a growing body of research in this area. Prior studies have investigated the experiences of nail salon workers through measures of chemical exposure assessment, symptom prevalence, and intervention evaluation. This research brings a distinct perspective, examining how the emotional, cultural, familial, and aspirational dimensions of workers’ lives shape their relationship to occupational health and safety. As we amplify these voices into the limelight, our findings highlight a recurring tendency for workers to describe individual-driven improvements rather than structural or systemic reform. This suggests that the conversation about nail salon workers’ health and safety must expand beyond workers themselves and involve the community beyond the salon.

We identified family as a core component of nail salon work, which may provide leverage for occupational health interventions. We found that nail salons are often family-owned businesses where relatives, extended family members, and children of nail salon workers come together, functioning as community hubs and workplaces. Furthermore, our participants described joining the nail salon workforce to provide better outcomes for the next generation. Existing literature that describes family in these settings has largely focused on the exposure risks to pregnant women, rather than on family as a motivation for salon work or as a potential point of intervention for improving occupational health [14, 16]. Our finding regarding intergenerational sacrifice is consistent with the broader understanding of the pathways of first-generation immigrants and their families [29]. However, intergenerational sacrifice has been underexplored in the occupational context of the nail salon. Given that interventions have been designed around personal safety concerns, there is a promising strategy: engaging the children of nail salon workers in efforts to improve salon health. Workers may be more receptive to concerns raised by their own children, for whom they are making sacrifices. Recent work indicates that second-generation Vietnamese-American children of nail salon workers share both the pride and burden associated with their parents’ work [30], suggesting they could be strong advocates. This strategy would be particularly important for families whose children intend to remain in the nail salon industry. However, the framework of sacrifice may also limit workers’ willingness to change salon conditions, as they see risks as an inherent part of the work and something to endure for their children.

The concept of job control emerged in our study. We observed the uncertainty of work hours, the competition among technicians and between salons, and the influence of customer preferences. Together, these factors may contribute to workers believing that they lacked the power to control their circumstances and could only control their responses to the demands of nail salon work. This idea is consistent with occupational health models where high demands and low control create job strain [31, 32]. Furthermore, our participants’ limited horizon on industry-level change can be reframed as a rational response to constrained work conditions. They are beyond the scope for individual workers or owners to implement. Despite many nail salons being family owned, nail salon workers lack a sense of empowerment to alter their work environment. These findings underscore the need for interventions to extend beyond individual behavior change approaches to a broader engagement with the family and community.

Customers emerged as powerful influencers of nail salon dynamics in our study, influencing salon practices through their expectations, preferences, and salon choice. Prior studies have found that customers can reinforce or undermine worker safety practices [6], yet customers have largely been absent from interventions. Thus, another promising approach is to include customers as stakeholders and to develop public-facing messaging that highlights protective practices and safe salon conditions. Indeed, such approach has precedent in California’s Healthy Nail Salon Recognition Program, where customers were willing to pay more at salons designated as a “Healthy Nail Salon” [7]. Indeed, one of our participants observed that customers were concerned about UV exposure from nail polish curing lamps, providing support for involvement of customers as stakeholders. Engaging customers could also help the cost barrier identified in prior nail salon intervention research [24, 25], linking safer practices to increased revenue.

Our findings point to two interventions that depart from the traditional education model. First, workers’ perceived lack of agency suggests a potential role for community liaisons who could serve as trusted intermediaries between salons and state cosmetology and health boards. Community liaisons could answer questions, communicate requirements, and reduce the fear of these agencies. Second, the pride and emotional component of nail salon work may themselves be a window of opportunity for intervention. Participatory approaches that invite nail salon workers to represent their own work, such as visual storytelling methods [21], can bring out the artistic component that workers value and also serve as an outlet for dispelling the emotional demands of the job. Such intervention may contribute positively to workers’ psychological and occupational health and bring together the nail salon community in a way that individually-targeted educational programs cannot.

One limitation of our study includes limited generalizability. Because our sample consisted of only Vietnamese nail salon workers in the Baltimore metropolitan area, our study may not reflect the experience of workers from other ethnic backgrounds in the nail salon workforce, such as Korean, Chinese, Nepali, and Hispanic workers [3], or nail salon workers outside of Maryland, as local cultures and regulatory landscapes may differ. Our study also shares limitations common in qualitative research. The use of a snowball sampling approach to recruit participants can introduce selection bias toward a sample that is more homogeneous. However, our findings are consistent with prior research in Pennsylvania and California, and with other groups. This suggests that nail salon workers of other backgrounds and in other areas share similar experiences and have navigated similar economic pressures and barriers as those in our sample, lending credence to the broader applicability of our findings.

In summary, our study provides a foundation for occupational health research and intervention development for nail salon workers in Maryland. The need for community-based interventions and participatory programs is shown in the priority consistently expressed by participants that family (not individual) success matters most to them. Our participants’ perceived lack of control further supports that the broader nail salon community needs to be engaged in any occupational health improvement efforts. Finally, engaging customers as potential partners in incentivizing investments in healthy products and practices holds significant promise. We look forward to community-wide, collaborative approaches that engage multiple stakeholders to uplift the nail salon worker population here and beyond.

## Data Availability

All data produced in the present study are available upon reasonable request to the authors.

## Acknowledgements

We thank the participants for sharing their experiences. We are grateful to the Maryland Vietnamese Mutual Association and Thao Bui for their collaboration on this project. We thank Dr. Tran Huynh for helpful discussions.

## Author contributions

Conceptualization: Kevin K. Nguyen and Emily M. Agree; Methodology: Kevin K. Nguyen and Emily M. Agree; Formal analysis and investigation: Kevin K. Nguyen; Writing–original draft preparation: Kevin K. Nguyen; Writing–review and editing: Kevin K. Nguyen and Emily M. Agree; Funding acquisition: Kevin K. Nguyen; Supervision: Emily M. Agree.

